# Development and content validity of the Clinical Needs Assessment for Myalgic Encephalomyelitis (CNAME)

**DOI:** 10.1101/2025.10.27.25338891

**Authors:** Sarah F Tyson, Russell Fleming, Keith J Geraghty, Peter Gladwell

**Author notes:** Corresponding Author: Prof Tyson.

## Abstract

**Purpose:** To co-produce the Clinical Needs Assessment for Myalgic Encephalomyelitis (CNAME) with people with ME and clinicians, and to assess its content validity.

**Methods:** Guided by the COSMIN guidelines, a draft CNAME was devised from relevant literature and lived experience. It was revised following piloting and cognitive interviews with the advisory groups and then completed online by people with ME. Content and face validity were assessed via deductive framework analysis of participants’ feedback. Construct validity (in terms of relevance, duplication and comprehensibility) were assessed using frequencies, cross-tabulations and response rates respectively. As the CNAME produces dichotomous data which are not summated and is not intended for repeated use further psychometric analyses are not possible or relevant.

**Results:** Four hundred people with ME participated. Comprehensibility was excellent (>97.5% item completion rate). There were no floor effects, but several items had a ceiling effect and were removed. Eight items demonstrated duplication and were combined. Participant feedback was positive, confirming that the CNAME addressed the issues that were important and relevant to them and was easy to complete.

**Conclusions:** The Clinical Needs Assessment for ME is a valid, feasible and acceptable measure of people with ME’s clinical needs.

## Introduction

Myalgic Encephalomyelitis (ME), also known as Chronic Fatigue Syndrome (CFS) is a complex multisystem condition of uncertain aetiology. The World Health Organisation classify it as a neurological disease, although immunological, metabolic, endocrine, and autonomic dysfunction are also widely reported^1,2^. Epidemiological studies are sparse, but most recent estimates indicate a prevalence of ~0.6% of the UK population, ie. ~410,000 people^3^. This is likely an underestimate however, as 45% of 1.9 million people in the UK with long covid are thought to meet the diagnostic criteria for ME/CFS^4^. ME/CFS affects women 1.5-2 times as frequently as mem, and all ages and ethnicities^5^. It is a highly disabling disease. Less than a third of people with ME/CFS are able to work^6^ and a quarter are severely or very severely affected, leaving them housebound, or even bedbound^7^. Consequently, the quality of life for people with ME/ CFS is poor and lower than many other disabling chronic conditions^8^. The economic impact of such a common and disabling condition is huge, estimated to be £3.3 billion/year in the UK, which is 50% higher than other chronic disabling conditions such as multiple sclerosis and lupus^9,10.^

There are no recognised curative or disease modifying treatments for ME/CFS, so clinical guidelines focus on holistic assessment to rule out alternative diagnoses and a patient-centred self-management approach to manage symptoms, activity levels and fluctuations^11^. A key element of clinical assessment is to identify patients’ ‘needs’. Needs being the issues that are important to the patient that they wish to address with the clinical service. A ‘needs assessment’ is the process of collecting information to identify patients’ needs.

Although, a new concept in the ME/CFS community, the notion of needs assessment is well established in other areas of health and social care, to the extent that the UK Care Quality Commission lists assessment of patients’ needs as a core competency for health care professionals and clinical services^12^. Establishing patients’ needs is a precursor to discussion and joint-decision-making to identify patients’ goals and priorities, and to prepare a care and support plan. The evidence-base regarding the impact of needs assessments is small, however evaluations indicate a positive effect in terms of patient and staff/service-related outcomes^13–18^.

Over 15 years ago, a systematic review broadly identified people with ME/CFS’s clinical support needs^19^. These were the need for respect and empathy from professionals, and help to:

- make sense of symptoms
- adjust views and priorities
- develop strategies to manage impairments, activity limitations and social participation
- obtain information regarding ME/CFS

However, these needs have not been operationalised into a format that could inform clinical care. Thus, the aim of this study was to work with people with ME/CFS and clinicians working in ME/CFS specialist services to co-produce a comprehensive checklist of people with ME/CFS’ clinical needs (called the Clinical Needs Assessment for ME, CNAME). This was part of a larger project to co-produce a clinical assessment toolkit for ME/CFS to support comprehensive patient assessment and care planning. As well as the CNAME, the toolkit contains assessments of ME symptoms, post-exertional malaise, activity limitations, and patient experience which will be published in due course.

## Method

The COSMIN methodology for assessing the content validity of patient reported outcomes was used to guide the study methods^20^.

### Context

This is a patient-led research study. Three of the authors have lived experience of ME/CFS and the fourth has extensive relevant clinical experience. They co-produced the CNAME with people with ME/CFS and clinicians working in ME/CFS specialist services in the UK. Two advisory groups provided their expertise by lived experience at every stage of the project. The ME advisory group consisted of people with ME/CFS who volunteered following publicity via UK’s ME Association’s newsletter, purposively sampled to ensure a wide range of age, geographical location, duration, and severity of ME/CFS were represented. An advisory group of clinicians working in specialist ME/CFS services was also convened via publicity through the British Association of ME Clinicians (BACME) networks, purposively sampled to ensure that a range of professions, experience working with ME/CFS, and types of ME/CFS specialist service were represented. Both groups contributed to all stages of the project.

As people with ME/CFS have limited energy and need to pace their activity carefully to avoid an exacerbation of symptoms and activity limitations (known as post-exertional malaise), it was a priority to minimise the ‘demand burden’ of contributing for the authors, ME advisory group and participants^21,22^. To achieve this, we consulted with the 25% ME group (a charity supporting people with severe ME/CFS) and advisory group members about how to minimise that burden. In both our interaction with the advisory group and data collection processes, we followed guidance to facilitate people with ME/CFS’s participation in research^23^ and the British Dyslexia Style Guide^24^ to maximise accessibility. We also offered a choice of formats for group members/ participants to respond. Most provided input by email (for group members) or online survey (for study participants). However, accommodations (such as paper copies, or completion by phone or video call) were also offered and participants were encouraged to contact the lead author to discuss any other accommodations.

### Construct and conceptual framework

As detailed above, clinical needs were defined as the issues the patient wishes to address with the ME/CFS service. Drachler et al’s^15^ systematic review of people with ME/CFS’s needs provided the conceptual framework. Our aim was to produce an assessment which enabled people with ME/CFS and clinicians to identify patients’ needs. This required a profile detailing the nature of the patients’ needs rather than a summated final score. Conceptually a summated score, such as the number or urgency/importance of the needs is not a useful construct to measure here. For example, someone with mild ME/CFS with concerns about maintaining or returning to employment would have a completely different needs profile to someone with severe ME/CFS who would be unable to work but may need help to maintain independence in the activities of daily living. However, they could have a similar ‘needs score’ if the number of needs were summated, or they were asked about the urgency/importance of their needs.

Thus, the CNAME was intended for use in clinical practice as part of the assessment process, rather than an outcome measure, or to monitor change over time.

### Target population

The CNAME was intended for use for anyone with ME/CFS. Thus our selection criteria were broad: People who had been diagnosed with ME/CFS in Great Britan.

### CNAME development: Item generation, response format, and recall period

An initial item list of ME/CFS clinical needs was generated from Draschler et al’s systematic review^15^ and expanded by the authors and the advisory groups drawing on their lived experience. We opted for a ‘yes/no/not relevant’ response format in which participants agreed or disagreed with statements regarding their need for each aspect of service provision (e.g. “I need mobility aids” – agree (score 1)/ disagree (score 0)/ not relevant (score 0)). We did not specify a recall period but used the present tense, i.e. we asked whether they had the need ‘now’ rather than at some time in the past.

### Evaluation of relevance, comprehensibility and comprehensiveness

The item list with response format was transferred to an online survey using Qualtrics Survey Tool and an introductory section and completion instructions added. As detailed above, we took extensive measures to maximise accessibility and minimise the completion burden of the survey. The relevance, comprehensibility and comprehensiveness were evaluated in two stages: piloting and cognitive interviews with the advisory groups, and surveying people with ME/CFS.

#### A) Piloting and cognitive interviews

The ME advisory group, and the clinicians’ advisory group both completed the pilot CNAME via electronic survey. Cognitive interviews were completed by email as this was the preferred format for members. They were asked to comment on, and make suggestions for changes regarding:

- the comprehensibility of the items, introductory section and completion instructions
- relevance of the items
- whether there was any duplication between items
- the comprehensiveness of the items and whether anything should be added
- ease of completing the survey and whether any further accommodations were needed.

As the information sought was quite specific, deductive framework analysis was used to analyse the advisory groups’ response using the broad codes of: relevance/ duplication; comprehensiveness; comprehensibility/ease of use; and accommodations, which were organised into sub-categories as necessary. Internal member checking ensured representativeness/internal validity.

#### B) Online Survey

To further assess the content validity, an online survey was undertaken. People who were diagnosed with ME/CFS in Great Britan were invited to complete the draft CNAME online. Recruitment was via publicity in ME/CFS support groups, the 25% ME group, and ME Association’s social media and publicity channels. Accommodations were offered for people who were unable to complete the survey online as detailed above.

As well as completing the item list of clinical needs, free text sections were added where participants were invited to add further comments regarding their needs, and feedback about the questionnaire, and how it might be improved.

The free text sections were analysed using deductive framework analysis as described above. A quantitative analysis was also used to assess relevance, duplication and comprehensibility:

- To assess relevance of the items, the frequency with which each need was identified was calculated. Items which were identified as a need by less than 25% of participants were considered candidates for removal.
- To assess duplication, cross-tabulations of items were undertaken. Cross-tabulations were used as inter-item correlations are not possible with dichotomous data. Items that showed >90% agreement (answering either ‘yes’ or ‘no’) were deemed to be duplicating the items’ construct and combined.
- Completion rates were a measure of comprehensibility. Items with a completion rate of <90% were considered for revision or removal.

As the checklist format produced unsummed dichotomous data, further psychometric evaluation of criterion or construct validity was not possible. Also, as the CNAME is intended for ‘one-off’ use during clinical assessment, reliability, sensitivity and responsiveness were not assessed as it was not intended to be used repeatedly.

## Results

Fifteen people with ME/CFS formed the ME advisory group. Ten were women, four were men and one was non-binary. Mean age was 53.4 years (sd 10 years) and their mean duration of ME/CFS was 15.3 years (sd 11.1 years). Most (n=10, 66%) were moderately affected (50-60% disabled); two were mildly affected (20-30% disabled) and two were severely affected (80-90% disabled). Nine professionals joined the clinicians’ advisory group. Two were men and two had personal lived experience of ME/CFS. On average, members had 3.6 (sd 3.4, range 1-12 years) years’ experience of working in a specialist ME/CFS service. The services they worked in were all clinic/out-patient (rather than in-patient) based. All offered remote consultations via phone or video conference for those who could not attend clinic and four (44%) also provided home visits. All took referrals for patients who were severe or very severely affected. One treated children as well as adults, and three (33%) accepted referrals for people with long covid as well as ME/CFS.

Four hundred people with ME/CFS completed the online survey. Three hundred and fifty-six (89%) were women, 41 (10%) were men and three identified as non-binary/other. Mean age was 53.5 (sd 13.11) years, and mean duration of ME/CFS was 14.9 (sd 11.4) years. Most participants reported their ME/CFS was at Level 3 (i.e. limited independence, n=201, 50%). However, 74 (18.5%) were severely or very severely affected (i.e. mostly housebound or bedbound respectively).

As a result of the cognitive interviews with the advisory group, the items were re-arranged into three sections: ‘the staff and service approach’, ‘understanding ME/CFS’, and ‘care and support plans’ which covered impairments/symptoms, activity/disabilities and participation. Several items were also added which addressed:

- social isolation
- employment support
- disability benefits
- explaining ME/CFS to others
- advocacy
- managing co-morbidities and their interaction with ME/CFS
- arrangements for ongoing support following discharge.

Minor changes to the wording and presentation were also suggested and incorporated. This produced an assessment with 42 items.

### Relevance of the items

All items were identified as a need by over 25% of participants, indicating no floor effects. The least frequently identified needs were the need for support with communication and socialising/ leisure, which were both identified by 42% of participants. However, several items demonstrated a ceiling effect, in that over 90% answered ‘agree’ to the need (Table 1).

**Table 1:**
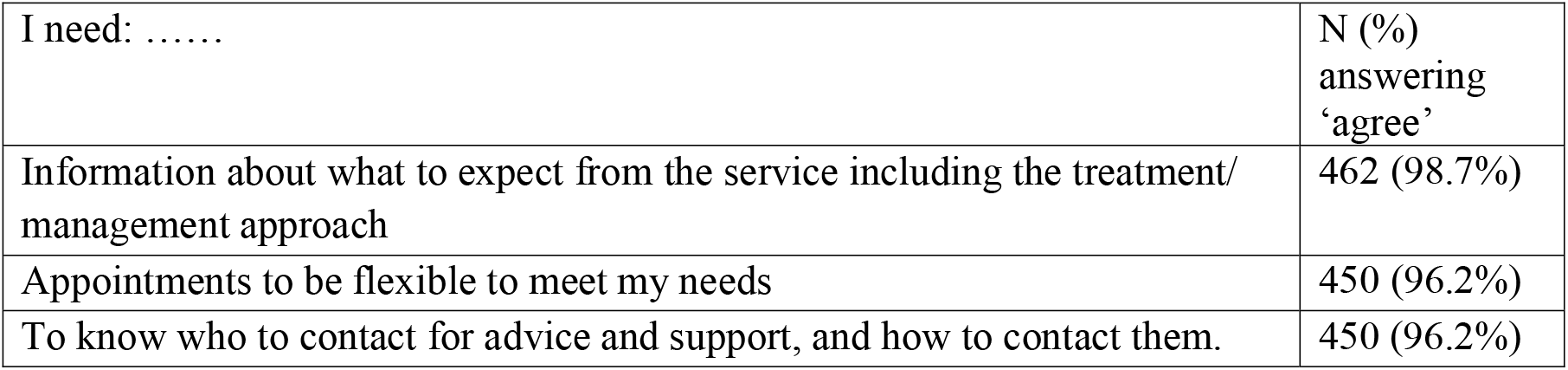
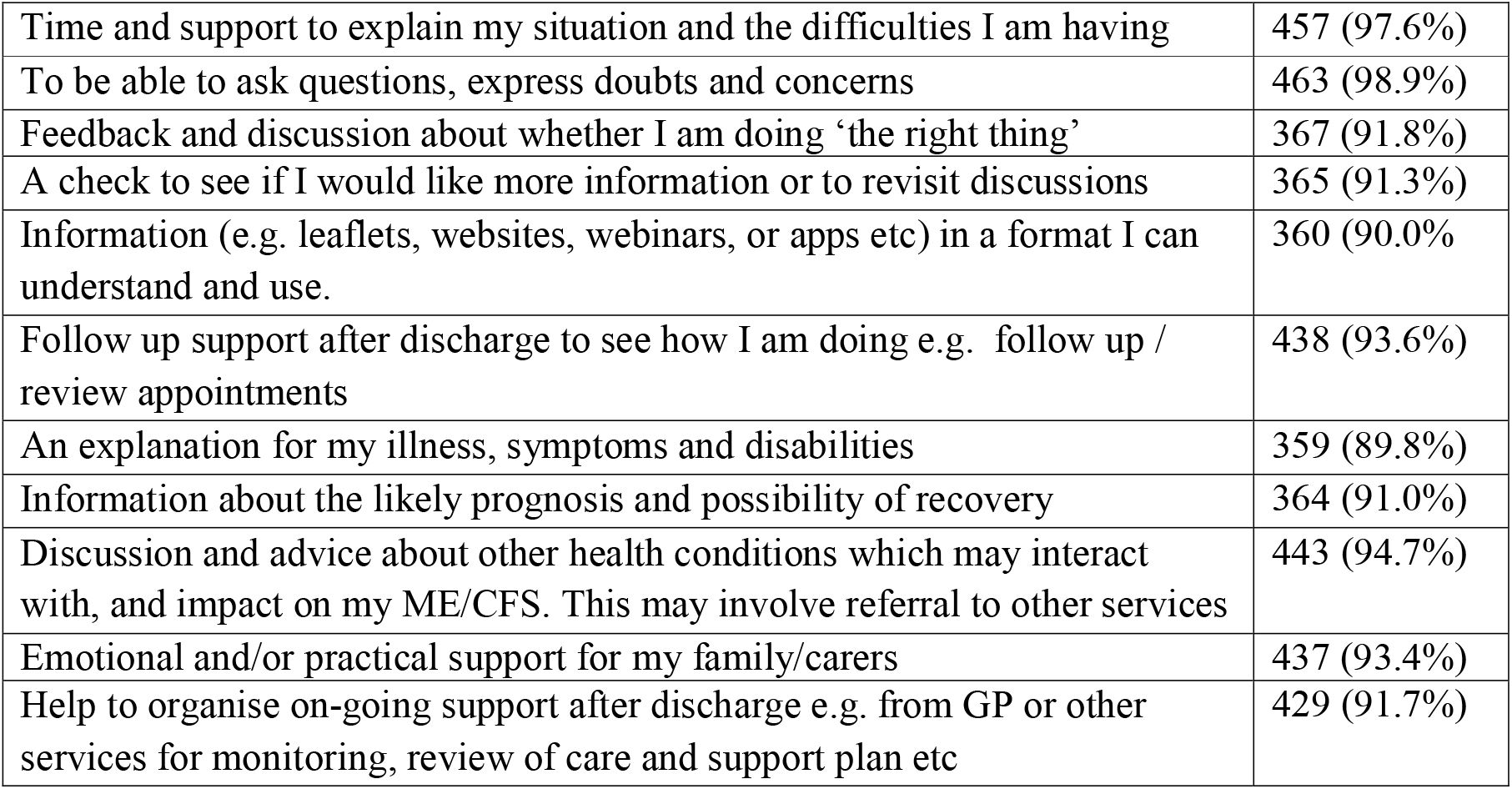
showing the items demonstrating a ceiling effect.

Consequently, these items were removed to minimise the completion burden for people with ME/CFS. However, as these issues are very important to people with ME/CFS, we did not want them to be unacknowledged. An introductory section was added to the assessment tool which explained that these needs were universal and important, and that services were expected to explicitly address them.

### Duplication of items

Eight items demonstrated high positive cross-tabulations (>90%) with other items, indicating duplication. These were combined into three overarching items as follows:

- Overarching Item 1: Advice and support on how to deal with the following symptoms:
  - physical fatigue
  - sleep disturbance
  - musculo-skeletal symptoms.

These items were combined, and reference to specific symptoms removed as these details would be provided in other assessments in the toolkit. The resultant item was *I need help to recognise and manage my symptoms*

- Overarching Item 2: I need help and support to:
  - identify and manage my activity limitation’s
  - avoid going past my energy limits or overdoing it
  - identify and manage fluctuations, setbacks and relapses.

These were combined into a single item; *I need help and support to identify activity limitations, manage them by pacing, and cope with fluctuations/relapses*.

- Overarching Item 3: I need help and support to
  - work out my needs and priorities
  - set my own goals.

These were combined into a single item: *I need help and support to work out my needs and priorities and set my own goals*.

#### Comprehensibility of items

Completion rates were very high at over 97.5% for all items, so none needed to be revised.

#### Participant feedback

Feedback from survey participants was positive, with participants noting the items were relevant and comprehensive, and the format was easy to complete. Several expressed relief that their reality had been captured, and their needs had been understood, as illustrated by Participant 305.

> *“…… it is clear that this work is being done by people who properly understand ME and its impact. When there is so much written that is inaccurate, misguided …*.. *It is actually a joy to know that someone, somewhere understands. Thank you”*

Many pointed out that they had had ME/CFS for a long time and support from specialist services was limited or, in many areas of Great Britain, non-existent. Consequently, they had worked out how to manage their ME/CFS and meet their needs as best they could themselves. This influenced their answers, and many noted that someone who was ‘new to ME/CFS’ would be likely to have a higher degree of need.

> *I have been ill for years so I have had to work a lot out by myself so I don’t really think a service could help me much now. If I had access to a service when I was first ill, my answers would be very different (Participant 267)*.

A few (n=11, 2.75%) found the response format (agree/disagree/not relevant) restricting and would have preferred a greater range of response options and to be able to add more detail. This was discussed with the advisory groups, who concluded that participants may have not fully understood that the purpose of the assessment was to highlight areas of greatest need to focus discussion with clinicians, rather than to record all the detail. They were also concerned that expanding the response options would add to the energy demand of completing the CNAME. Thus, we resolved to add further explanation about the purpose of the assessment and how it would be used in the introductory section, rather than to change the response format, as eloquently illustrated by Participant 201.

> *I hope the intention is that the questionnaire would be followed by an in-depth discussion. People often don’t know what it is that they don’t know. Especially early on, they don’t know what help they need or what topics they need to know about*.

A free text box was also added at the end of the assessment for those who wished to add further detail to their assessment. Two people reported that they found the questionnaire too long, and two found it tiring to complete. Several noted that they had had assistance to complete it, which had been encouraged if participants found it taxing to complete on their own.

As a result of the revisions made, the final version of the CNAME had 21 items.

## Discussion

The results of this study demonstrate that the Clinical Needs Assessment for ME (CNAME) is a valid assessment tool. By using a co-production approach with people with ME/CFS and clinicians in ME/CFS specialist services and following gold standard guidance regarding assessment development,^20^ we are confident the CNAME has content validity in that it represents the issues that are important and relevant to people with ME/CFS and clinical services in an accessible format. Other aspects of psychometric evaluation are not possible because the CNAME produces a profile rather than summated data, or not relevant (as it is not intended for repeated use).

Consequently, we recommend the CNAME to enable people with ME/CFS and their clinical teams to identify and better understand patients’ needs as part of the clinical assessment process. This can contribute to goal-setting and prioritisation processes and inform care planning and management strategies. However, the implementation of needs assessments raises some important issues. In the CNAME, we described clinical needs as *“issues that are important to you* [the patient with ME/CFS] *and a priority for you to address as part of your care and support plan”*. However, a service’s ability to do this depends not only on the patient’s presentation and priorities, but also on the supply of, and effectiveness of care, i.e. one cannot benefit from care that is either unavailable or ineffective. ME/CFS specialist services are known to be sparse in Great Britain and many areas have no provision at all^25^. Existing services are also very varied in terms of their remit and scope, staffing levels and skill mix and may not be resourced to address all the needs identified in the CNAME^25^. For example, few services have dietetic input so the ability to address nutritional needs are limited^25^. Anecdotally, some services would deal with this by developing relationships with other services to support referrals, while others would consider it beyond their remit. There is an ethical and pragmatic dilemma about whether to ask patients about needs if the resources to address them are unavailable. Not only could this raise unachievable hopes and expectations but also could give an inaccurate picture of how effectively the service was working within the resources available to them. There is an argument that clinical needs assessments could be individualised to the services’ provision, i.e. to only ask about needs the service feels able to meet. However, we advise against this, as it would limit the CNAME’s scope to support service benchmarking and improvement. We urge services which are not resourced to meet some of the CNAME’s needs to investigate what resources are available from other services locally, so that patients can be advised and referred onwards, and/or to work with managers and service commissioners to develop services to address those unmet needs.

To the authors’ knowledge, this is the first tool to assess people with ME/CFS’ needs, so comparisons with other assessments are not possible. The main strength of this study lies in its large sample size, co-production with people with ME/CFS and clinicians and rigorous adherence to best practice research reporting oguidance. The main limitation lies in the representativeness of the sample. Our sample was predominantly middle-aged women with prolonged lived experience who are moderately affected. This is consistent with other large cross-sectional surveys of people with ME/CFS^3,9,26–28^ so we are confident the sample is representative. However, the recruitment methods may have biased towards English speakers who are active on social media and in ME organisations.

Health records regarding ME/CFS are notoriously inaccurate, incomplete and inconsistent^3,9,28,29^ which influenced our recruitment methods and the data collected. We recruited from outside health services because a) they rarely have sufficiently accurate records to effectively identify people with ME/CFS, and b) we wanted to involve all people with ME/CFS rather than the very small minority who have access to services. For the same reason, confirmation of a ME/CFS diagnosis was not sought. This does mean we may have recruited some people who would prove to have other diagnoses if thoroughly assessed or accurately recorded.

Finally, nearly 20% of our sample reported that they were severely or very severely affected (that is, essentially housebound or bedbound). This is slightly lower than the 25% who are estimated to be severely affected in prevalence studies^3,9^. This likely reflects the proportion of people who were too ill to complete the survey, even with the accommodations offered.

Further research is needed to better understand the demographics of ME/CFS; to accurately diagnose and record it; and to investigate the needs of underserved groups.

In conclusion, the Clinical Needs Assessment for ME is a valid, feasible and acceptable measure of people with ME/CFS’ clinical needs. The final version of the CNAME is found in the supplemental material and is freely available for others to use.

## Supporting information

Supplemental File 1. Final version of the CNAME

## Data Availability

All data produced in the present study are available upon reasonable request to the authors

## Acknowledgments

The authors would like to thank the ME and clinicians’ advisory groups who co-produced the CNAME.

