## Supplemental File 1. Final version of the CNAME for "Development and content validity of the Clinical Needs Assessment for Myalgic Encephalomyelitis (CNAME)"

This questionnaire is a checklist to identify the issues that are important to you and a priority to address as part of your treatment/management plan. These are known as 'clinical needs'.

It has been developed by people with ME/CFS and clinicians working in specialist ME/CFS services and is intended to be used with the other assessment tools in the ME Association's Clinical Assessment Toolkit.

The CNAME takes about 10 mins to complete in one go, but you can take as long as you want to complete it.  If you need help from another person, or another person to complete it on your behalf, that is fine. If you would prefer to complete it by phone, need any other adjustments or have any other questions, please contact the person who sent this form.

There are 22 questions, arranged in two sections: 'Understanding ME/CFS' and 'Care and Support Plan'. Each question has a statement, 'I need ….' And the answer format asks you to agree (that you do have that need) or to disagree (that you do not have that need), or that the need is not important or relevant to you. There are also some open questions that enable you to add for detail if you wish.

### CLINICAL NEEDS ASSESSMENT FOR ME

| | |
|---|---|
| Name: | NHS Number |
| Date: | No of assessment |

Please note that during the development of the CNAME, several aspects of ME/CFS were noted to be so important that everyone highlighted them as a 'need'. So, they have not been included in this final version to minimise the effort of completing the form. However, you should be aware of them and can expect the service to provide the following:

- information about the service. For example, what to expect, contact details, and arrangements for support after discharge
- information about ME/CFS in a user-friendly format.
- flexibility to meet your needs. However, please note this may be limited by the resources available within the service.
- staff to be approachable and knowledgeable and to give you the time you need.
- support for your family/carers

For other issues, please answer the following questions by indicating whether you need help by 'agreeing' or 'disagreeing'.

| **I need ……** | **Agree** | **Disagree / Not relevant** |
|---|---|---|
| **SECTION 1: Understanding ME/CFS** | | |
| Accommodations to attend the service | | |
| Help to explain ME/CFS and my difficulties to others | | |
| Help to explain ME/CFS and my difficulties to 'officials' | | |
| Support to involve my family and others in my care | | |
| Support to build my confidence to manage my illness | | |
| Opportunity to 'meet' others with ME/CFS. | | |
| Advice about keeping in touch with others, managing/maintaining relationships, or breaking out of social isolation | | |
| Information about ME/CFS support groups/ charities or other resources | | |
| **Section 2: Care and Support Plan** | | |
| Help to work out my needs and priorities and set my own goals | | |
| Support to develop an action plan to address my needs and priorities | | |
| Help to recognise and manage my symptoms | | |
| Information and support about medications | | |

| |
|---|
| Help to identify and manage my activity limitations including pacing, fluctuations and relapses |
| Help with basic activities of daily |
| Help with domestic/ home care |
| Help with family life |
| Help with work/ education |
| Help with benefits and social care |
| Help with mobility and ‘getting around’ |
| Help with socialising and leisure |
| Help with communication |
| **Comments** |
| Do you need any accommodations or adjustments to attend the service? (Please detail) |
| FREE TEXT |
| Is there anything else you would like us to know about your needs? |
| FREE TEXT |
